# Analytical evaluation and critical appraisal of early commercial SARS-CoV-2 immunoassays for routine use in a diagnostic laboratory

**DOI:** 10.1101/2020.10.20.20215970

**Authors:** Amanda Cramer, Nigel Goodman, Timothy Cross, Vanya Gant, Magdalena Dziadzio

## Abstract

**BACKGROUND:** The objective of this study was to evaluate the performance characteristics of early commercial SARS-CoV-2 antibody assays in mild and asymptomatic subjects to enable the selection of suitable serological assays for routine diagnostic use within HCA Healthcare UK.

**METHODS:** We used serum samples from a pre-Covid era patient cohort (n=50, pre-December 2019), designated SARS-CoV-2 negative, and serum samples from a SARS-CoV-2 RT-PCR-positive cohort (n=90) taken > 14 days post symptom onset (April-May 2020). We evaluated 6 ELISA assays including one confirmation assay to investigate antibody specificity. We also evaluated one point-of-care lateral flow device and one high throughput electrochemiluminescence immunoassay.

**RESULTS:** The ELISA specificities ranged from 84-100%, with sensitivities ranging from 75.3-90.0%. The LFIA showed 100% specificity and 80% sensitivity using smaller sample numbers. The Roche CLIA immunoassay showed 100% specificity and 90.7% sensitivity. When used in conjunction, the Euroimmun nucleocapsid (NC) and spike-1 (S1) IgG ELISA assays had a sensitivity of 95.6%. The confirmation IgG assay showed 92.6% of samples tested contained both NC and S1 antibodies, 32.7% had NC, S1 and S2 and 0% had either S1 or S2 only.

**CONCLUSIONS:** These first generation assays were not calibrated against reference material and the results are reported qualitatively. The Roche assay and the Euroimmun NC and S1 assays had the best sensitivity overall in our hands. Combining the assays detecting NC and S1/S2 antibody increased diagnostic yield. A portfolio of next generation SARS-CoV-2 immunoassays will be necessary in any future studies of herd and vaccine induced immunity.

## Introduction

The current worldwide coronavirus pandemic caused by severe acute respiratory syndrome coronavirus 2 (SARS-CoV-2) has resulted in millions of confirmed infections and cases of the associated COVID-19 disease.^1^

The clinical manifestations of acute infection vary widely from asymptomatic disease to severe viral pneumonia and lung failure^2,3^ and the extent of subsequent chronic organ damage and COVID-19 disease remains to be established. A definitive diagnosis of SARS-CoV-2 infection currently relies on the use of RT-PCR to identify the virus in respiratory samples ^4-6^. This only identifies current illness or a viral carriage. The performance of the RT-PCR test is dependent on various factors including time of sampling, viral load and how thoroughly the sample is taken from the nasopharynx; its estimated sensitivity varies and falls around 80-85%.^7^ Consequently, a significant proportion of infected individuals may be missed from screening programmes.^8^ In contrast, robust serology assays which reliably detect the presence of antibodies against SARS-CoV-2, can determine whether individuals with or without symptoms have previously been infected, thus providing valuable information about prior exposure for epidemiological purposes and the individual patient.^9,10^

The introduction of serological testing to HCA Healthcare UK was rapidly implemented in April 2020 to cover a variety of scenarios ahead of National Policy. A strategic decision was taken to roll out a healthcare worker screening programme for purposes of staff and patient protection, and to have a robust assay in place in anticipation of evolving national or local guidance. Accordingly, we scanned the first-generation antibody assay horizon for candidate tests we could rollout into routine laboratory diagnostics.

During the early months of the pandemic, numerous SARS-CoV-2 immunoassays were rapidly developed and placed on the market. These assays used different antigenic proteins; some used whole virus lysate, recombinant nucleocapsid (NC) or full spike (S) proteins, while some use modified proteins or peptides of the NC or specific domains of the S protein - glycoprotein 1 (S1), spike glycoprotein 2 (S2) or RBD (receptor-binding domain).

In this study we initially evaluated the analytical performance of CE marked IgG ELISAs and a lateral flow device, selected based on the availability in the UK at the time. We later compared these to the analytical performance of the Roche “total antibody by electrochemiluminescence (CLIA) immunoassay”, chosen because the platform for the Roche assay (the Cobas system) is already in place at HCA Laboratories.

## Materials and methods

### Sample set

In total, 140 patient serum samples were obtained from the Serology and Immunology Departments within HCA laboratories during March and April 2020.

#### Specificity cohort

50 pre-COVID era samples taken before December 2019 were collected from the Serology Department, kept frozen at -20^°^C.

#### Sensitivity cohort

10 serum samples from adult hospitalised patients with confirmed SARS-CoV-2 infection (days post symptoms: mean 21.2 days, range 15 - 33) were obtained from excess serum samples sent to the Serology Department in March 2020.

A further 80 serum samples from healthcare workers (HCW) employed by HCA and proven to have had SARS-CoV-2 infection by viral nucleic acid detection (RT-PCR) from an upper respiratory tract (nasopharyngeal) swab were tested in an accredited laboratory.

Serum was taken from these individuals after their return to work and after two negative swab tests by RT-PCR, having isolated for 2 weeks after their confirmed RT-PCR positive result. Staff were asked to report the date of onset of Covid-19 associated symptoms. In all cases, sensitivity cohort samples were taken > 14 days post symptom onset to optimise detection of SARS-CoV-2 antibodies. Seventeen (17%) subjects were asymptomatic and sixty-three (63%) had mild symptoms.

The study was an audit of routine sera and was reviewed by the local HCA Audit and Research Committee. All samples were anonymised, and written consent was obtained from the HCW individuals for the audit. Consent is not required for surplus samples used for assay verification.

## Methods

### Lateral flow immunoassay (LFIA)

The LFIA was only evaluated for specificity using 20 of the pre-COVID era samples due to the limited number of kits to hand. For sensitivity, 80 of the HCW samples were tested by staff at HCA Primary Care facilities.

We used the COVID-19 IgG/IgM Rapid Test (CTK Biotech Inc, Poway, USA), performed in accordance with the manufacturer’s instructions. The test was accepted if the ‘C’ line (control line) showed the appearance of a coloured band, with the result designated as either positive (G and/or M band development) or negative (C line only).

### Enzyme-linked immunosorbent assays (ELISA)

We assessed 5 qualitative ELISAs and one confirmatory ELISA from 4 different manufacturers as outlined below. All ELISA assays were run using the automated DS2 Analyser (Dynex® Technologies, Inc).

Due to the speed these assays were brought to market, they were not calibrated against any reference material (e.g. IRP 67/86 of human serum immunoglobulins) or international standards. ELISA results were qualitative and based on a calculated ratio (of the relative antibody concentrations) set by the manufacturer or a “confidence index” assigned by the manufacturer.

#### Epitope Diagnostics

The EDI^™^ Novel Coronavirus COVID-19 IgG ELISA kit (KT-1032, Epitope Diagnostics, San Diego, USA) utilises a recombinant full length nucleocapsid protein to measure human anti-SARS-CoV-2 IgG antibody in serum. The assay was run as per manufacturer’s instructions and the results were calculated as recommended.

#### Euroimmun

The Anti-SARS-CoV-2 NCP ELISA (IgG) and Anti-SARS CoV-2 S1 (IgG) ELISAs (Euroimmun Medizinische Labordiagnostika, Lübeck, Germany; EI2606-9620-2G and EI2606-9601G respectively) were performed according to manufacturer’s instructions.

The NC assay wells are coated with modified nucleocapsid (NC) of SARS-CoV-2 which according to the manufacturer contains “diagnostically relevant epitopes”, whereas the S1 assay wells are coated with an S1 domain of the spike protein of SARS-CoV-2 expressed in the human cell line HEK 293.

#### Dia.Pro

The Dia.Pro COVID-19 IgG ELISA and the COVID-19 IgG Confirmation ELISA kits (Diagnostic Bioprobes Srl, Milan, Italy) are intended for use in conjunction as an initial screen, and subsequent confirmatory assay, respectively, for antibodies to SARS-CoV-2. The Dia.Pro COVID-19 IgG ELISA contains a mix of recombinant “immunodominant antigens” (nucleocapsid, S1 and S2) whereas the confirmation test is composed of individual wells coated with these antigens and hence enables to determine antibody specificity against NC, S1 and S2 present in the patient’s sample.

#### NovaLisa®

NovaLisa® SARS-CoV-2 9 (Covid19) IgG ELISA (obtained from NovaTec Immunodiagnostica Gmbh, Dietzenbach, Germany, product code COVG0940) utilises microtiter plates coated with SARS-CoV-2 antigens (antigen names are not provided).

### Chemiluminescent assay (CLIA)

The Roche Elecsys total antibody (IgG and IgM) assay was run on the Cobas® e801 analyser. The Elecsys Anti-SARS-CoV-2 assay is an electrochemiluminescence sandwich immunoassay for qualitative detection of both IgG and IgM in human serum and plasma against a recombinant SARS-CoV-2 nucleocapsid antigen.

## Statistical analysis

For each method, sensitivity, specificity and 95% confidence intervals were calculated using the R statistical computing environment^11^. A pairwise comparison was performed between each unique combination of assays using a two-sample Z-test for equality of proportions. A p-value of ≤0.01 was used as a threshold for significance to correct for multiple testing.

## Results

### Sensitivities and specificities

Four commercial anti-SARS-CoV-2 IgG ELISA kits were evaluated initially, all of which measured antibodies raised against the recombinant nucleocapsid (NC) antigen of the virus. Subsequently, the Euroimmun Anti-SARS-CoV-2 IgG S1 ELISA, which measures antibodies to the S1 (S) domain of the spike protein, became available and was included in the evaluation.

Table 1 shows sensitivities and specificities of these 5 ELISA assays and the LFIA and Roche CLIA assay. To maximise specificity and sensitivity for each ELISA, borderline results were classified as negative for the EDI and DIA.PRO kits, and positive for the Euroimmun and NOVALisa kits.

**Table1:** Sensitivities and specificities of ELISA, LFIA and CLiA assays for SARS-CoV-2 antibody detection shown with 95% confidence intervals (CI)

| ASSAY | Number of Serum Samples |  |  |  | Sensitivity<br>(95% CI) | Specificity<br>(95% CI) |
| --- | --- | --- | --- | --- | --- | --- |
|  | RT-PCR confirmed cases |  | Pre-COVID control |  |  |  |
|  | True Positive | False Negative | True Negative | False Positive |  |  |
| **ELISA** |  |  |  |  |  |  |
| EDI | 67 | 14 | 49 | 1 | 82.7(72.4-89.9) | 98.0 (88.0-99.9) |
| DIA.PRO | 83 | 7 | 42 | 8 | 92.2(84.1-96.5) | 84.0(70.3-92.4) |
| NOVALisa | 67 | 22 | 50 | 0 | 75.3(64.8-83.5) | 100.0(91.1-100) |
| Euroimmun NC | 81 | 9 | 50 | 0 | 90.0(81.4-95.0) | 100.0(91.1-100) |
| Euroimmun S1 | 80 | 10 | 50 | 0 | 88.9(80.1-94.3) | 100.0(91.1-100) |
| **LFIA** |  |  |  |  |  |  |
| CTK | 62 | 16 | 20 | 0 | 79.5(68.5-87.5) | 100.0(80.0-100) |
| **CLIA** |  |  |  |  |  |  |
| Roche | 68 | 7 | 25 | 0 | 90.7(81.1-95.8) | 100.0(83.4-100) |

The 5 ELISA assays had specificities ranging from 84-100% and sensitivities ranging from 75.3-90.0%%. The LFIA showed 100% specificity and 79.5% sensitivity, although the small sample sizes should be noted. The Roche CLIA immunoassay showed 100% specificity and 90.7% sensitivity.

The Roche assay and the Euroimmun NC and S1 assays had the best sensitivity overall, all with 100% specificity and when combined, the Euroimmun NC and S1 assays had a sensitivity of 95.6%. The Euroimmun ELISAs against 2 viral proteins were therefore chosen as the ELISA(s) most appropriate for deployment.

### Euroimmun assays

The sample cohorts were classified as hospital, mild disease and asymptomatic according to details given at time of sampling. The results of the Euroimmun NC and S1 are shown in Table 2 when grouped according to these classifications.

**Table 2:** Percentages of patient cohorts with antibodies to nucleocapsid (NC) and S1 (S) by Euroimmun ELISA kits

| **Patient Cohort** | **Negative**<br>**(NC and S1)** | **NC** | **S1** | **NC or S1** |
| --- | --- | --- | --- | --- |
| **Hospital (N=10)** | 0.0% | 90.0% | 100.0% | 100.0% |
| **Mild (N=63)** | 4.8% | 88.9% | 87.3% | 95.2% |
| **Asymptomatic**<br>**(N=17)** | 5.9% | 94.1% | 88.2% | 94.1% |

These results show that patients who had severe disease all had either S1 or NC antibodies however the mild/asymptomatic cohort showed little difference between the groupings. Of the HCW cohort, 17/80 (21.3%) were asymptomatic.

### Dia.Pro Confirmation assay

The Dia.Pro conformation assay was performed to identify the specific antibodies present in the samples from RT-PCR positive patients that tested positive by the Dia.Pro ELISA screen assay. These results are shown in table 3. In total, 52 samples were tested on the confirmation assay.

**Table 3:**
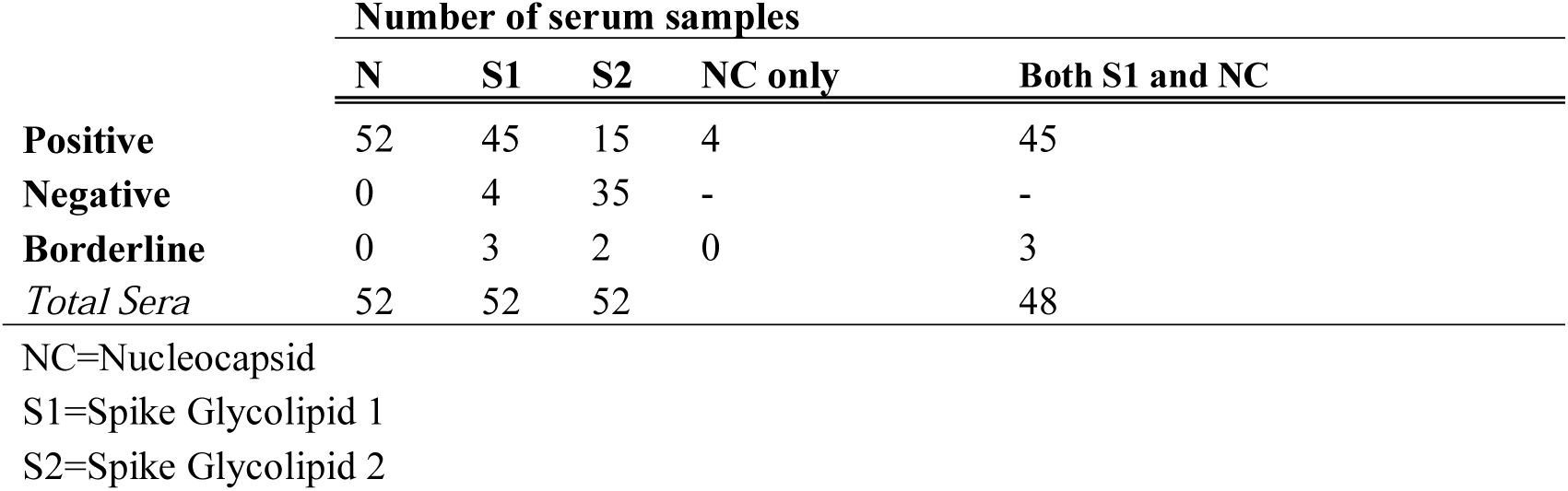
Number of samples showing different specificities of antibodies to the major SARS-CoV-2 antigens using the Dia.Pro Confirmation assay

|  | Number of serum samples |  |  |  |  |
| --- | --- | --- | --- | --- | --- |
|  | N | S1 | S2 | NC only | Both S1 and NC |
| **Positive** | 52 | 45 | 15 | 4 | 45 |
| **Negative** | 0 | 4 | 35 | - | - |
| **Borderline** | 0 | 3 | 2 | 0 | 3 |
| *Total Sera* | 52 | 52 | 52 |  | 48 |

NC=Nucleocapsid
S1=Spike Glycolipid 1
S2=Spike Glycolipid 2

All 52 samples contained NC antibodies and the number of samples that had all three antibodies was 17/52 (32.7%). 48/52 (92.6%) contained both NC and S1 antibodies. Of the hospitalised patients, only 1/10 (10%) had anti-NC IgG only and 4/10 (40%) had all three specificities (anti-NC, anti-S1 and anti-S2 IgG). None of the 52 samples contained S2 antibodies only.

### Comparison between the assays: sensitivity and specificity

Sensitivity: pairwise comparison showed a statistically significant difference between NovaLisa and Dia.Pro ELISA screen assay, p<0.005.

Specificity: pairwise comparison showed a statistically significant difference between NovaLisa and Diapro ELISA screen assay, p<0.001, Euroimmun S1 and Diapro ELISA screen assay p< 0.001, Euroimmun NC and Diapro ELISA screen, p<0.001 and NovaLisa and Diapro ELISA screen, p<0.001.

### Comparison of discordant samples

Of the confirmed positive RT-PCR samples from the healthcare workers that were run across all 7 platforms, contradictory results were found in 23/75 samples as shown in table 4. Each of these 23 samples were positive on either of the two Euroimmun assays.

**Table 4:** Discordant samples across all seven platforms

| **Sample ID** | **CTK** | **DIAPRO** | **EDI** | **Euroimmun Nc** | **Euroimmun S1** | **NOVALisa** | **Roche** |
| --- | --- | --- | --- | --- | --- | --- | --- |
| 61 | Pos | Pos | Pos | Neg | Pos | Pos | Neg |
| 62 | Neg | Pos | Pos | Pos | Neg | Pos | Pos |
| 64 | Neg | Pos | Pos | Pos | Pos | Neg | Pos |
| 65 | Pos | Neg | Neg | Pos | Pos | Neg | Pos |
| 66 | Neg | Neg | Neg | Pos | Pos | Neg | Pos |
| 67 | Neg | Pos | Pos | Pos | Neg | Pos | Pos |
| 69 | Pos | Neg | Neg | Neg | Pos | Neg | Neg |
| 70 | Neg | Neg | Neg | Neg | Pos | Neg | Neg |
| 71 | Neg | Pos | Neg | Pos | Neg | Neg | Pos |
| 72 | Pos | Pos | Pos | Pos | Pos | Neg | Pos |
| 73 | Pos | Pos | Neg | Pos | Pos | Neg | Pos |
| 78 | Pos | Pos | Neg | Neg | Pos | Neg | Neg |
| 82 | Pos | Pos | Pos | Pos | Pos | Neg | Pos |
| 83 | Pos | Pos | Pos | Pos | Pos | Neg | Pos |
| 91 | Neg | Pos | Pos | Pos | Neg | Neg | Pos |
| 98 | Pos | Pos | Pos | Pos | Pos | Neg | Pos |
| 107 | Pos | Pos | Neg | Pos | Pos | Neg | Pos |
| 112 | Neg | Pos | Pos | Pos | Pos | Pos | Pos |
| 113 | Neg | Pos | Neg | Pos | Pos | Neg | Pos |
| 114 | Pos | Pos | Pos | Pos | Pos | Neg | Pos |
| 117 | Pos | Pos | Neg | Pos | Pos | Neg | Pos |
| 118 | Pos | Pos | Neg | Pos | Pos | Pos | Pos |
| 131 | Neg | Pos | Pos | Pos | Neg | Pos | Pos |

It was noted that 3/80 (4%) of the confirmed positive RT-PCR samples from healthcare workers were negative across all the SARS-CoV-2 antibody tests performed in this study.

## Discussion

We evaluated analytical performance characteristics of five commercial ELISAs, one POC rapid test and the Roche CLIA immunoassay, in order to find a suitable assay to make SARS-CoV-2 antibody testing available for routine diagnostic use for HCA Healthcare UK’s COVID19 screening and diagnostic strategy. Our paper relates to an analytical evaluation and from the immunological perspective; it does not imply that all subjects with positive RT-PCR for SARS-CoV-2 will make virus specific antibodies. Immunological mechanisms of humoral response to SARS-CoV-2 infection are still unclear and underlying conditions, e.g. antibody deficiency, are not considered in this evaluation.

We used a cohort of pre-COVID19 era samples to assess specificity and a cohort of HCA healthcare workers to assess and evaluate the analytical sensitivity of serological assays in mild and asymptomatic disease.

We found that the ELISA assay with the highest sensitivity was the Euroimmun NC and S1 kits (90% and 88.9% respectively) and when used in combination, this increased sensitivity to 95.6%.

The Roche Elecsys was found to have 100% (95%CI 83.4-100) specificity and 90.7% (95%CI 81.1-95.8) sensitivity. These results were in accordance with another evaluation, which found this assay had a specificity of 100% (95%Cl 99.1-100) and sensitivity of 86.1% (95% Cl 76.5-92.8) for samples taken ≥14 days from symptom onset and 86.7% (95%CI 76.8-93.4) for samples taken ≥21 days since symptom onset.^12^

The Cobas® analyser is a fully automated high throughput platform. This platform, established in our laboratories, potentially offered considerable logistic advantages to enable rapid rollout of antibody testing not only for our staff, but also to ensure capacity for high volumes of sample requests for SARS-CoV-2 antibodies in our patients. These elements of performance and surge capacity prompted our decisions to routinely choose the Roche assay.

We do not have the longitudinal antibody monitoring data to better inform the kinetics of the antibody responses in our cohort. Similarly, we were not able to establish negative and positive predictive values since the prevalence of SARS-CoV-2 infection is an estimate and was not known at the time of the study. Four percent of RT-PCR positive subjects had no detectable antibodies to SARS-CoV-2 suggesting that not all otherwise healthy individuals will mount humoral response to the virus. This has been noted by Shields et al.^13^ and several research groups.

We noted significant differences in terms of assays sensitivity and specificity (NovaLisa assay was the least sensitive (75%) and Dia.Pro was the least specific (84%) but the most sensitive (92%)). There was significant variability between the assays with regards to the individual sample result (positive/negative) as shown in Table 4. These differences can be explained by several analytical variances between the assays, for example antigen type and source and its immunogenicity. The antibody assays we evaluated used a variety of different viral protein antigens, e.g. recombinant modified or full-length protein, mixed antigens or antigens not specified (NovaLisa). It is well known that SARS-CoV-2 proteins have complex 3D structure and complex antigens purification process which is a hurdle for the manufacturers who attempt a fast development of new immunoassays for the emerging disease.^14^.

Potential assay interferences may lead to false positive immunoassay results. For example, the Dia.Pro kit assay insert states that “10% of the reactive normal population collected before the outbreak showed reactivity to nucleocapsid”. It remains to be established whether this positivity in the Dia.Pro assay which uses “immunodominant recombinant antigens” is due to cross-reactivity with other Coronaviridae or it is due to the unknown interferences or cross-reactions with the components in the sample. In our hands, Dia.Pro had lowest specificity, calculated at 84%, of all the ELISA assays we evaluated.

The sensitivity and specificity of these assays, in our hands, differed from those reported by the manufacturers. These early assays came on the market following a quick assay evaluation using very small or small sample size coming mainly from hospitalised individuals with severe disease known to make strong antibody response^15^ and therefore not optimised for population based serological sampling and investigations of asymptomatic or mildly symptomatic individuals with SARS-CoV-2 infection. Hence, the assay sensitivities and specificities figures established by the manufacturers were higher than those established by us and other UK diagnostic laboratories.

The assays results were reported qualitatively and expressed in ratios and arbitrary units and not as concentrations. The positive cut-offs for the assays were defined by the patient population chosen by the manufacturer which typically was a group of older hospitalised patients with severe disease. Ideally, one would perform a ROC curve on each assay using the same data set which would limit the variability of the results observed from the different methods.

These early assays were not calibrated against any reference material, unlike the newer assays, e.g. ELIA Phadia SARS-CoV-2 IgG assays, that use the IgG calibrators traceable to the International Reference Preparation (IRP) 67/86 of Human Immunoglobulins. The quality of the SARS-CoV-2 antibody assays should improve with the 1^st^ WHO international standard which is expected in December 2020. External quality assurance schemes have now been established to allow for better quality control of these assays.

It is important to keep in mind that the major use of SARS-CoV-2 serology assays was to be for screening in populations with low disease prevalence and for determining the prevalence of SARS-CoV-2 exposure in asymptomatic and individuals with mild disease. CDC has recommended to combine independent assays to improve positive predictive values^16^. This highlights the importance of our data with comparison of different assays and our findings of best sensitivity and specificity of SARS-CoV-2 antibody assay by combining NC and S1 Euroimmun assays.

The selection of an appropriate antibody assay and the timing of the measurements are of paramount importance for laboratories and will further inform seroprevalence studies as well as clinical outcomes^17^. In the recent systematic review of 491 papers by Huang et al.^18^, the most used assays were binding assays led by ELISA. Median time of detection of SARS-CoV-2 antibody was 11 days and antibody kinetics varied across the severity gradient, with antibodies remaining detectable longer after severe illness. There is a knowledge gap here since antibody kinetics data come from the symptomatic individuals and not from subclinical or asymptomatic groups.

Assays based on detection of antibodies to nucleocapsid were attractive for commercial companies since these antibodies are detected sooner post-infection. However, they do not last at detectable levels in mild disease and they have potential for cross-reactivity with other coronaviruses. However, spike proteins are the main immunodominant antigens involved in virus receptor binding (S1) and cell membrane fusion (S2) and anti-spike protein antibodies are better correlates of viral neutralisation. Therefore, anti-SARS-CoV-2 vaccines target spike protein to elicit immune responses^2, 17,18^.

Recent evidence suggests that deep profiling of the acute humoral immune response to SARS-CoV-2 may play a role in predicting disease outcomes. Atyeo et al.^19^ found that spike-specific antibody responses in hospitalised patients with SARS-CoV-2 infection were associated with convalescence and better outcomes while functional antibody responses to the nucleocapsid were elevated in deceased individuals. In that study, a combination of five SARS-CoV-2 specific antibody measurements enabled the authors to distinguish individuals with different disease trajectories. Hence, the diverging immune response and the signature with a higher spike: nucleocapsid ratio appears to be a powerful biomarker in disease activity and in predicting clinical outcomes.

In summary, we have shown our experience in evaluating and implementing fit-for-purpose SARS-CoV-2 assays for rollout in our routine laboratory diagnostics. Our study demonstrated that the platforms tested have similar analytical performance (except for the Dia.Pro assay which in our hands had low specificity). Our results show that diagnostic laboratories like us can provide data that will inform future diagnostic, prevention and treatment pathways and in this way can play a role in fighting the pandemic. However, we also showed that no single assay will be sufficient to dissect antibody response in COVID19 disease. A portfolio of new generation robust antibody assays is necessary to facilitate further understanding of COVID19 disease and its prevention, similarly to the existing antibody assays for viral hepatitis. This portfolio should allow the detection of high quality neutralising antibodies produced as a result of the germinal centre response as well as assays specific for the individual viral proteins that distinguish between the immunity due to natural infection, vaccination and recrudescence.

## Data Availability

The data referred to in the manuscript is held securely within the department of Immunology at HCA laboratories.

## Disclosure statement

The authors have no conflicts of interest to disclose.

## Funding information

No external funding was required for this study.

## Notes

### Competing Interest Statement

The authors have declared no competing interest.

### Author Declarations

The study was reviewed by the local HCA Audit and Research Committee.

